# Experiences of mental health and wellbeing support for NHS staff during the COVID-19 pandemic: a reflexive thematic analysis

**DOI:** 10.1101/2022.06.15.22276446

**Authors:** Corinne Clarkson, Hannah R. Scott, Siobhan Hegarty, Emilia Souliou, Rupa Bhundia, Sam Gnanapragasam, Mary Jane Docherty, Rosalind Raine, Sharon A. M. Stevelink, Neil Greenberg, Matthew Hotopf, Simon Wessely, Ira Madan, Anne Marie Rafferty, Danielle Lamb

## Abstract

Staff in the National Health Service (NHS) have been placed under considerable strain during the COVID-19 pandemic; whilst NHS Trusts provide a variety of health and wellbeing support services, there has been little research investigating staff perceptions of these services. Moreover, the research that does exist typically includes only clinical staff, despite a large proportion of patient-facing NHS workers being in non-clinical roles. We interviewed forty-eight clinical and non-clinical healthcare workers from eighteen NHS Trusts in England about their experiences of workplace health and wellbeing support during the pandemic. Reflexive thematic analysis identified that perceived stigma around help-seeking, and staffing shortages due to wider socio-political contexts such as austerity, were barriers to using support services. Visible, caring leadership at all levels (CEO to line managers), peer support, easily accessible services, and clear communication about support offers were enablers. Our evidence suggests Trusts should have active strategies to improve help-seeking. This could involve providing all staff with regular reminders about support options, in a variety of formats (e.g. email, posters, mentioned in meetings), and easily remembered single points of access, delivered by a mix of in-house and externally-provided services, to cater for those more and less concerned about stigma and confidentiality. In addition, managers at all levels should be trained and supported to feel confident to speak about mental health with staff, with formal peer support facilitated by building in time for this during working hours. As others have pointed out, this will require long-term strategic planning to address workforce shortages.

## Introduction

Amidst the COVID-19 pandemic, the challenge of providing care for huge numbers of sick and dying patients has placed immense strain on National Health Service (NHS) staff in England, coming, as it did, after a decade of austerity (Charlesworth et al., 2021). While there is evidence of sharp increases in symptoms of anxiety and depression in the general population, compared to pre-pandemic levels (Daly & Robinson, 2021), there is conflicting evidence about whether healthcare workers (HCWs) are showing higher prevalence of mental disorders than the public (Kwong et al., 2020; Murphy et al., 2020; Paul et al., 2021), although we do know that NHS workers feel unwell due to pressure of work, and feel unsupported. For example, the 2021 NHS Staff Survey found that in the previous 12 months, 44% of staff felt unwell due to work stress (up from 40% in 2019), that 13% did not feel safe speaking up about concerns at work, and 40% were not confident that their organisation would address any concerns (NHS England, 2021).

There is currently limited research on the support needs of HCWs, including their views on what is desirable and effective, when and how it should be provided, and what prevents and enables uptake (Billings, Seif, et al., 2021). There is a particular paucity of research on the needs of non-clinical staff (both patient-facing, e.g. porters, receptionists, and non-patient-facing, e.g. finance and administrative personnel) who make up a significant proportion (47%) of the English NHS workforce (NHS Digital, 2020). Throughout the pandemic, new interventions and services have been put in place alongside existing ones to support HCWs, such as relaxation rooms, mental health and practical helplines, free parking, and externally provided Employee Assistance Programmes. However, evaluation of such interventions has been scarce (Billings, Seif, et al., 2021) and so organisations are at risk of investing in new and existing services that have limited evidence bases, using resources that could be better employed elsewhere.

Qualitative research is particularly valuable in understanding what potential service users do and do not want, as well as how services may or may not be accessed, the context in which they are used, and how beneficial users find them. A review of qualitative work during the current pandemic and previous viral outbreaks internationally found that HCWs are commonly challenged by high workloads, limited resources, and communication issues at work, with mixed views about the extent of how adequate the support received is (Billings, Ching, et al., 2021).

There is evidence that frontline HCWs in the UK faced similar difficulties with lack of clear and accessible messaging from their organisations, limited consultation from management, and barriers to using practical and psychological support during the COVID-19 pandemic (Billings, Seif, et al., 2021). Participants in such studies frequently report a reliance on peer support; whilst this can be an effective protector against negative wellbeing outcomes in times of stress (Hu et al., 2012), it may also indicate limitations in formal workplace support offerings. However, non-clinical HCWs are typically under-represented in such studies, and so very little is known about their preferences or needs. In addition, while exploration of structural, systemic, and individual barriers to accessing psychosocial support has been recommended by others in this field (Billings, Seif, et al., 2021; Liberati et al., 2021), this has not been the focus of much recent work.

To address these gaps in the existing evidence base, the current study explored the experiences of clinical and non-clinical HCWs in England of support services at their workplace, with a specific focus on barriers and enablers to meeting their mental health and wellbeing (MHW) needs during the COVID-19 pandemic.

## Methods

Ethical approval for this study was granted by the Health Research Authority (reference: 20/HRA/210, IRAS: 282686) along with local Trust Research and Development approval.

### Participants

This study was nested within ‘NHS CHECK’, a longitudinal cohort study monitoring the physical and mental health of English HCWs during the COVID-19 pandemic (Lamb et al., 2021). Eighteen NHS Trusts (acute and mental health) from across England were invited to participate in the study, purposively selected to represent a variety of geographic locations and serving populations with a range of sociodemographic characteristics in urban, suburban, and rural contexts. All staff in each participating Trust were emailed invitations to take part in the study, which initially involved completing an online survey taking approximately 10-20 minutes, and collected data on: sociodemographic characteristics; occupational roles and settings; support services they were aware of/had used/found helpful; and a range of validated psychosocial measures (the primary outcome measure was the General Health Questionnaire, GHQ; Goldberg et al., 1997). Participants for this study were individuals from the cohort who had agreed to be contacted about further research, and were purposively sampled to ensure representation across age, sex, ethnicity, job role, as well as use of support services offered.

### Recruitment

Recruitment to the NHS CHECK study occurred from April 2020 to January 2021. NHS CHECK participants who had agreed to be contacted about further research were emailed invitations to the study with an information sheet, between April 2021 and August 2021. Participants who responded with interest were sent links to an online consent form and an online calendar to book into an interview slot at a time of their choosing.

### Procedure

Interviews were conducted via phone or MS Teams with one of four interviewers (CC, HRS, SH and ES). A semi-structured interview schedule was developed and piloted with NHS CHECK Advisory Board members, who suggested minor changes around language used. The schedule covered use of and access to support services available within the workplace, perceived benefits and drawbacks to services, and alternative sources of support. Minor changes to questions or phrasing of questions were made to the schedule depending on whether a participant was aware of any available support services, and whether they had used any of these services (Appendix 1).

Interviews were audio recorded and transcribed verbatim by a transcription service, with identifying information removed, de-specified or pseudo-anonymised. All interviewers spent a brief period before each interview building rapport with each interviewee, and gave each participant a chance to reflect on their interview once it had been completed. Interviewers made clear to participants that they were academic researchers (though some have previous clinical experience), independent from the NHS or participants’ Trusts, and that any data used from interviews would be anonymised such that no individual, group, or workplace would be identifiable.

Participants who completed the interview received a £25 gift voucher in recognition of time volunteered for the study.

### Analysis

Demographic data were derived from the primary NHS CHECK dataset. To analyse interview data, we followed the six recursive stages of reflexive thematic analysis developed and detailed by Braun and Clarke (Braun & Clarke, 2006, 2021). Nvivo 12 software was used for analysis. Coding and theme development was carried out collaboratively, with authors initially independently coding transcripts, and refining code clusters and then themes through discussion. An inductive coding process was followed, developing an initial coding framework which was reviewed by other NHS and mental health and wellbeing (MHW) support staff to improve external validity. A more deductive process was followed thereafter, with refinement of the framework carried out through discussion as more data were coded.

Reflexive practice was followed throughout the data collection and analysis period, with interviewers individually reflecting on their subjectivity and discussing this with other authors. Some members of the authorship team are or have worked as clinicians within the NHS, and so are closely positioned to the topic and participants in this article; one interviewer (CC) previously practiced as a clinician, the others (HRS, SH and ES) have academic backgrounds. The lead author (CC) was particularly aware of her own strong feelings about the pressures of working in the NHS previously, and took steps such as keeping reflective notes and discussing with clinical colleagues the effects of hearing other NHS staff talk about their own distress.

## Results

Forty-eight participants from eighteen NHS England Trusts were interviewed for the study. Key characteristics of the sample are shown in table 1, with services used by the participants listed in table 2.

**Table 1:** Age, sex, and role characteristics of sample, and knowledge/use of services

| Characteristic |  | N (%) |
| --- | --- | --- |
| Age |  | Mean = 43; SD = 10.95 |
| Sex | Male | 20 (42) |
|  | Female | 28 (58) |
| Primary role | Clinical | 25 (52) |
|  | Non-clinical | 23 (48) |
| Knowledge of services | Aware of at least one available support service | 48 (100) |
|  | Unaware of any available support services | 0 (0) |
| Use of services | Ever used one or more support service | 21 (44) |
|  | Never used any support service | 27 (56) |
| Psychological distress | Met GHQ<sup>a</sup> cut-off score ($\geq 4$) | 26 (62) |

<sup>a</sup>GHQ: General Health Questionnaire.

**Table 2:** Workplace support services used by participants

| Type of support service (self-reported as used by participants) | Number of participants who used this service* (%) |
| --- | --- |
| Individual counselling | 13 (27) |
| Wellbeing smartphone applications | 4 (8) |
| Wellbeing hub | 4 (8) |
| Occupational therapy | 3 (6) |
| Mindfulness sessions | 2 (4) |
| Psychoeducational sessions | 2 (4) |
| Free food | 2 (4) |
| Q&A/ reflection sessions with management | 2 (4) |
| Free car parking | 1 (2) |

\*some participants used more than one service, and some may not have used any, so % do not add up to 100.

We identified three stacked organising themes, each influencing each other; 1) socio-political context, 2) organisational culture, 3) individual experience (Figure 1). Pseudonyms are used when quotes are given below. We assigned pseudonyms with the intention of reflecting the participants’ key demographics, while maintaining anonymity. Additional quotes illustrating each theme are available in Appendix 2.

**Figure 1.**
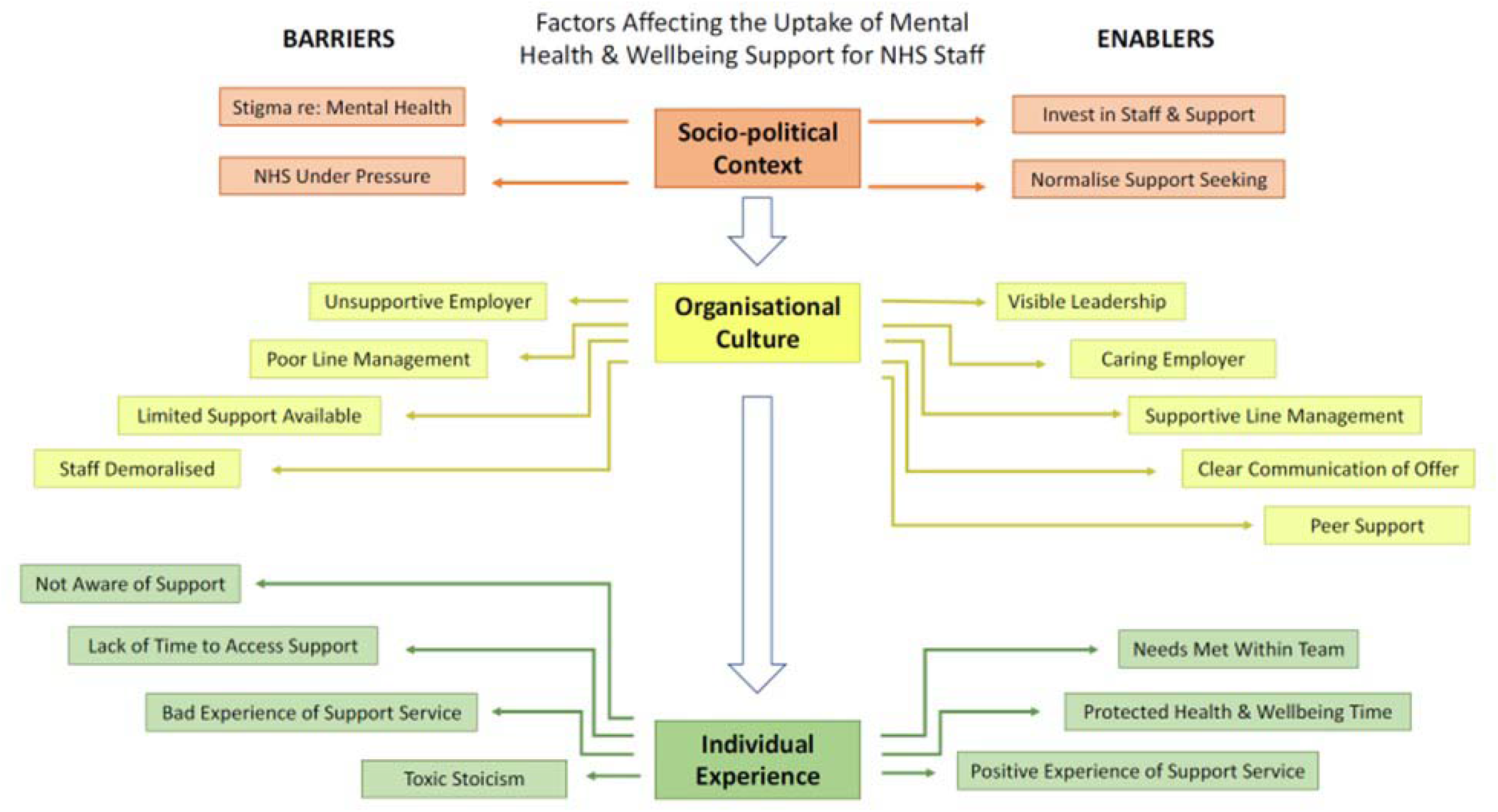
Thematic map

### Socio-political Context

Participants described systemic pressures on the NHS, including workforce shortages, and a perception of reduced real-terms funding while patient numbers and demand have increased. This was perceived to have created an NHS culture that is unable to prioritise staff wellbeing, and where staff struggle to provide a good service and take care of themselves. Extreme additional demands of the pandemic on pre-existing pressures contributed to conditions in which participants found it difficult to do their jobs well, and was experienced as detrimental to wellbeing.

> *“It was the huge pressures that we were under to meet performance criteria above our own wellbeing and seeing high caseloads. When you’ve got 70-80 patients on your caseload when really we should be having about 35, having to meet the demands of that job. And having to see more patients because colleagues are off because they had to shield, really struggling. And there was so much emphasis on the political stuff, it’s like a business. What about the patient?” Warren, psychologist*

Multiple participants suggested that sector-level investment in safe staffing levels would facilitate use of support services, allowing staff the time to look after themselves and therefore provide safe and effective care to patients.

Participants also described a perceived stigma in society about mental health, and how this stigma of mental health was a barrier to uptake of psychological support. For some, their professional role as helpers created a further barrier to accessing practical and in-person health-related support:

> *“People aren’t willing to talk about it because even within the Health Service there is such a stigma over mental health that there is still a reluctance, and a reluctance to actually admit that staff struggle sometimes*.*” Ah Lam, nurse*

Some participants described the pandemic-induced focus on staff wellbeing as having positive impacts on the expansion of existing support services for staff and introduction of new services. However, there was also concern that wider issues around remuneration were detrimental to wellbeing, particularly when compared to responses in other countries:

> *“It just seems like now we’re out of the public eye it’s gone back to ‘well you’re just a worker, you work for us, you are expected to be professional but it’s just you work for us, we pay you’, so that’s it end of. No matter what you’ve worked throughout the pandemic and how many lives you saved. With Scotland and Wales giving bonuses and increasing nurses pay as well it’s just another big slap in the face that we’ve worked just as hard, harder*.*” Clemmie, nurse*

### Organisational Culture

Participants had complex relationships with their employing Trust and wider NHS. They were acutely aware of strain on themselves and colleagues but often minimised their own needs while functioning in environments defined by high workload and pressure in a novel situation.

Some noted a lack of sustainability of support offerings, having used newly introduced services at the beginning of the pandemic, but having had them removed after the first wave. The same participant described losing a valued practical resource early in the pandemic:

> *“We had two fallout rooms that were quickly taken by management, and those fallout rooms were confidential spaces for staff to talk with another member of staff if they’re struggling, to have supervision in, to make a private call*.*” Caroline, support leadership*

Other participants highlighted that support services were historically and currently oversubscribed. Alternatives to investment in formal support services were received particularly poorly by participants, and were perceived as benefitting the image of Trusts rather than their staff:

> *“You have wellbeing champions, you have champions for safeguarding, you have a champion for drug and alcohol, you have a champion for this, a champion for that, but I don’t know it just feels a bit like ‘yes we’ve got a champion, and it looks good’, rather than the actual having depth and weight*.*” Jonathan, mental health and wellbeing support worker*

The way participants talked about their Trust varied, revealing a range of relationships with their employer, with perceptions ranging from very caring to exploitative. For example, enabling factors such as visible leadership within a caring employer, providing supportive line management, and clear communication of support offer created an environment protective of staff wellbeing where needs were met within teams.

Participants valued visible and responsive leaders who provided opportunities for their voices to be heard and experiences to be shared within the organisation. Leaders needed to understand the demands of frontline work and make staff health and wellbeing a Trust priority, with the majority of participants stating that line managers had an impact on their wellbeing:

> *“She sometimes has this knack of knowing us sometimes better than we know ourselves and if she sees that we’re slightly stressed or we look like we’re putting pressure on ourselves she will pull us to one side and say ‘are you OK, is everything OK?’. So as a group under her leadership we’ve all developed those skills*.*” Laura, physiotherapist*

Other participants had negative experiences of direct leadership, feeling that their line managers were not compassionate or person-centred in their interactions with staff and this became not only a barrier to accessing support but a source of distress. Some felt that their line managers could be a barrier to seeking help directly or via referral to support services where relationships with them were strained or distant. Some of these participants recognised that managers were often untrained and unsupported in their own roles, leaving them ill-equipped to support teams and practicing poor line management:

> *“If you step up to the role of a ward manager, it’s like ‘right yes you get some training in 12 months, in two years, whenever it becomes a chance’ because as soon as you step up to the role you are then just treading water and figuring out what it is you are doing*.*” Stephen, mental health clinician*

Senior leaders were seen as critical in determining a Trust’s culture. Participants expressed a sense of discomfort where they felt that organisational finances and metrics of success were prioritised over patient and staff needs, and this could contribute to a sense of feeling unsupported. Presenteeism, repurposing staff spaces, poor line management, and limited availability of support contributed to negative perceptions of the Trust and left staff feeling demoralised. Where Trusts were perceived as caring employers, providing a variety of accessible practical and psychological support services and promoting awareness and use of services, this was reassuring, even for staff who did not use them.

Clear communication of support offered by the Trust needed to happen through a variety of channels, with attention to the fact that not all staff have access to computers or time to read emails. Frequent reminders and memorable single points of contact from support service staff were beneficial in raising awareness and simplifying access to support services. Line managers were again seen as key to increasing awareness of an encouraging access to available services.

> *“Everybody needs that face-to-face conversation rather than something that is buried in an email or right at the back of a broadcast because people don’t read that stuff. Most people don’t read that stuff. People don’t notice posters on the walls, they walk past it, they see it, they don’t register it. Whereas like when we had the chaplaincy service when we had someone sitting there saying how are you today, if you need to chat about anything just let me know, here’s my number*.*” Petra, management*

The majority of participants valued informal peer support as immediately accessible and relevant to the unique needs of each team’s working environment, some suggesting that support from colleagues was more beneficial than formal services available from the Trust. Support occurred naturally between groups of colleagues, but was sometimes facilitated by managers.

> *“We all met in the park just to have a bit of a chat about how we were feeling, almost like a debrief, which our Trust or executive team suggested would be a good idea for teams to get together and talk about these things*.*” Ruksanah, NHS researcher*

A minority of participants felt that relationships with colleagues could be complex, and discussing mental health was not always possible nor desirable with peers, given perceived stigma.

### Individual Experience

Individual experiences of support were influenced by organisational culture and personal outlook. Limited awareness of support services, lack of time to use them, previous bad experiences of support services, and internalised toxic stoicism (participants feeling they had to do their jobs regardless of personal cost) as a response to poor working conditions, and demoralisation acted as barriers to uptake of mental health and wellbeing support.

All participants were aware of at least one existing support service, although awareness about the variety of support services available to them differed considerably between participants (with no clear pattern across Trusts). Some were highly aware of a range of resources available to them through their workplace and knew how to access services, whilst some only were aware of a relatively small number (typically occupational health or psychological support services) and were not clear on routes to access. Just under half of participants had used at least one support service; most of these participants felt that their experience was helpful and that it had made a positive difference in their ability to cope at work. Participants faced a mixture of practical and personal barriers to the use of support.

Some staff with relatively low awareness of services had little interest in seeking support within their workplace, having met their support needs externally through social support or professional services outside the workplace. However, there were others who may have benefitted from support but were not fully aware of what was available nor how to access it, suggesting they had not received effective communication from Trusts:

> *“I was just a bit lost on where and who should I go to. Like I say I saw about psychological services to help people during the COVID period kind of thing but I was unsure when I accessed them whether it was appropriate*.*” Chomba, nurse*

The most frequently cited barrier to uptake of MHW support services for individuals was simply lack of time to use them, particularly in clinical roles. Participants typically felt that their workload was too high to take time out of working hours to access support and were reluctant to use services outside of typical working hours, or on days off.

> *“There’s no time at work, so it has to be done out of hours. But out of hours means after 5pm which is not an option for most people or weekends. I don’t think it’s doable*.*” Ahmed, doctor*

Some Trusts were able to mitigate this barrier by facilitating access to services during protected time in working hours; this approach was widely suggested by participants, particularly clinicians, who did not have it implemented at their Trust.

Almost all participants felt overworked to some extent. For some, this led to feeling undervalued and as though they were unable to do their job well. It was common for staff to feel internal (personal) pressure to be resilient, particularly when comparing themselves to colleagues who appeared to be coping better. This feeling was characterised by low expectations, cynicism, reluctance to admit needing help, and ‘just getting on with it’. We have conceptualised such feelings as ‘toxic stoicism’, where the desire to fulfil caring responsibilities despite workers’ own feelings of distress led to potentially unhelpful presenteeism, and self-comparison with colleagues who appeared to be coping better. Such feelings created barriers to seeking support in a working environment that could feel damaging to health and wellbeing within roles that, particularly amongst clinicians, were integral to their identities.

> *“I think going up to your manager and saying I’m just really stressed out I just need to take some time out of the day maybe something has just overwhelmed you, you just get looked at like you are failing. You don’t want to let your patients down, you don’t want to let your colleagues down and that’s the way it’s kind of portrayed to us is you are the weak one. It’s not what you’ve been through it’s your fault if you can’t cope. Everybody else is coping well why can’t you*.*” Steve, nurse*

When staff did seek help, they sometimes had bad experiences of support services. Services at some Trusts were found to be unresponsive or unable to meet ongoing needs. Psychological services were received generally positively, but it was more common for participants to feel dissatisfied with these services if provided internally rather than externally. Of participants who used or tried to use internal occupational health, most were dissatisfied, having concerns about confidentiality and criticising long waiting times, lack of clarity about what support could be offered, and referral routes only via line managers.

> *“They referred me to occupational health to see whether I should do anything different. So I went to occupational health and said would anything be beneficial, what is my risk and she’s like ‘oh no you are fine you can go back to work*.*’ I was like ‘is there a reason why I was referred?’ She was going ‘oh your manager will tell you*.*’ I was like ‘OK*.*’ I couldn’t work out what the point of the conversation was. I think that’s the only thing I’ve ever had in 15 years of working for the NHS with occupational health*.*” Fiona, administrator*

In contrast, where NHS Trusts were perceived to have cultivated positive cultures prioritising staff welfare, investing in support services, and providing supportive line management, participants reported feeling cared for by their employer and this supportive environment could meet needs without participants feeling as though they required additional specific services. Factors perceived as curating positive experiences of support services included self-referral, rapid access to varied interventions including expert-led therapeutic support, and having the option to receive confidential support from somebody outside of the employing Trust:

> *“Continuing with external rather than provide it inhouse I think more people would be willing to take it up and even if we had to meet face to face off premises actually knowing you are away from the work area actually I think will make people feel much more willing to be open and honest and therefore get the support they need*.*” Kirsten, radiologist*

## Discussion

This study provides a detailed exploration of how NHS staff viewed the various mental health and wellbeing support services available to them over the pandemic. Overall, we found that study participants had reasonable awareness of the support services available at their workplace, although a need for improved communication was noted by a sizeable minority. Most of those who had used support services reported having had a positive experience, although responsiveness, capacity, and concern about confidentiality commonly impacted on experience. However, participants often reported facing a range of barriers and facilitators to help seeking; the most commonly reported as being important was the culture and leadership of the organisation, which could considerably help or hinder an individual’s ability to use services as well as their general feelings of level of support and care from their Trust. The majority of participants felt their workload was too great; resulting in limited time and energy to engage with support. Furthermore, feelings of toxic stoicism frequently impacted participants’ ability to use support services. Many of these underlying structural issues existed prior to the pandemic, but have been further compounded by increased pressures on the NHS.

### Research in context

#### Socio-political Context

Our findings align with research on stigma around mental health at the societal level: currently the subject of a 10-year national reduction programme (‘Time to Change’) showing small effect sizes with persistent geographic/demographic inequalities (Henderson et al., 2020). Studies focusing on healthcare cultures have similarly found that staff experiencing symptoms of mental health disorders can feel stigmatised (Knaak et al., 2017). NHS leaders describe staff reluctance to engage as a barrier to implementing support services (Quirk et al., 2018) while staff describe stigma and lack of time as barriers to engagement (Billings, Bloomfield, et al., 2021). However, stigma and failure to seek help has also been reported in many other occupational settings such as the military, emergency services and media professionals (Greenberg et al., 2009; Stevelink et al., 2020; Williamson et al., 2019). Where staff do feel able to speak to colleagues, this informal peer support is highly valued, as discussed below. Combining psychological and practical support has been found to increase uptake (Chen et al., 2020), and has been suggested to normalise support seeking through meeting physiological and wellbeing needs for NHS nurses (Maben & Bridges, 2020). To our knowledge, there has been no recent review of existing occupational health and wellbeing services in NHS Trusts, and it is unclear whether the health sector is ahead of or behind other sectors in terms of mental health and wellbeing support.

The resourcing issues raised by many of our participants, in particular staff shortages, are well known to NHS leaders, and to politicians, with recent reports of NHS vacancy rates at 1 in 10 full-time equivalents (NHS Digital, 2021). Strategic plans, including the 2019 NHS Long Term Plan (NHS, 2019) and the 2020/21 NHS People Plan (NHS, 2020), prioritise staff wellbeing and support but have been widely criticised for failing to address underlying causes of poor wellbeing without the means to address longstanding clinical workforce shortages (The Health Foundation, 2020; The Kings Fund, 2021). This, despite evidence that workforce shortages and staff burnout negatively affect patient outcomes (Aiken et al., 2014).

#### Organisational Culture

As suggested by participants in this study, NHS culture can be complex, and vary between, and even within, Trusts. Cultural change presents challenges but is widely accepted to drive performance and safety improvements (Mannion & Davies, 2018).

Leadership was described by participants as influencing their perception of the culture of their workplace and their capacity to use support. Generally, participants expressed the need for communicative and supportive leadership both within Trusts and across the NHS. This aligns with a body of evidence demonstrating that the perceived adequacy of training and support from employers for HCWs has a positive impact on psychological wellbeing (Baxter et al., 2021). The benefits of compassionate and collective leadership, from the highest level (e.g. CEOs) cascading down to direct line managers, further support the case for transforming NHS Trust leadership (West et al., 2014, 2017).

The relationships between supportive or poor line management, mental health and wellbeing of HCWs, and uptake of psychosocial support, remain understudied in current literature (Cabarkapa et al., 2020; Kisely et al., 2020; Preti et al., 2020). One rapid review discusses the protective impact of supportive teams (Kock et al., 2021), and a recent survey of 558 UK HCWs revealed strong desire for supportive and visible managers alongside holistic support offers from employers (Siddiqui et al., 2021). There are some existing interventions to support line managers to be more able to speak with staff about their mental health, however higher quality research on this topic is needed (Akhanemhe et al., 2021).

Other qualitative research of UK HCWs has similarly highlighted the tension between leadership, culture, and access to support, noting a gap between policy and practice around access to support (Vera et al., 2021) and the need for flexible and self-managed use of services (Billings, Seif, et al., 2021). Similarly, exploration of HCW experiences on the frontline during COVID-19 also found a desire for early support through line managers, and that managers need similar support to be able to offer this support (Newman et al., 2021). HCWs are not alone in reporting a need to supportive leadership, with similar findings in other organisational contexts (e.g. (Jones et al., 2012)

Generally positive experiences of peer support have been commonly found in research on HCWs (Billings, Ching, et al., 2021), and quantitative research offers evidence that it can be protective against psychological distress among HCWs during viral outbreaks (Cabarkapa et al., 2020).

#### Individual Experiences

Some participants had limited awareness of what available support services were for and how to access them; related research has found that clear communication was important in determining MHW outcomes amongst redeployed staff surveyed in one Trust (Walker & Gerakios, 2021). Our findings highlight the difficulties of defining ‘clear communication’, with many staff unaware of services that Trust administrative and communications teams presumably believe to be advertised clearly.

In line with findings of our study, lack of time and not catering to those who do shift-work (particularly night shifts) were known to be significant barriers to engagement with MHW support in pre-pandemic research (Billings, Biggs, et al., 2021; Brand et al., 2017; Braun & Clarke, 2019). Staff desire for protected time and flexible access to support is described in other qualitative studies, along with failure to provide it, contributing to the perception by staff of unsupportive employers who have employee mental health as a low organisational priority (Billings, Seif, et al., 2021).

In line with some previous qualitative studies, our participants reported feeling unvalued, reluctance to engage, getting on with it (Billings, Seif, et al., 2021). We have built on this work, aggregating these experiences as a unified concept in our analysis, ‘toxic stoicism’, which strongly resonated with staff when we discussed our interpretation of the data with NHS staff. There was considerable overlap between toxic stoicism and descriptions of understaffing, excessive workloads, low pay, and high attrition, indicating that this is a maladaptive response to existing resource pressures, which have been exacerbated by the pandemic.

### Strengths and limitations

The participants interviewed represent a diverse range of UK HCWs, with participants from a number of Trusts, ethnicities and staff roles, and a balance of ages and both men and women. Importantly, and building on previous similar research which focussed on specific staff groups (Billings, Seif, et al., 2021), we included all types of staff (clinical and non-clinical). Additionally, the sample comprised both help-seeking and non-help-seeking staff and so was able to investigate barriers and facilitators to accessing services from participants with a range of perceptions and experiences of services, including those not predisposed to use them. However, as this study was nested within a larger cohort study, participants were self-selected from an already self-selected sample and so were inclined to engage with research and willing to speak to people about their experience. To check whether we accessed staff coping with the highest levels of burden (or, conversely, unrepresentatively low levels of burden) we compared the proportion meeting General Health Questionnaire cut-off score in our sample to the full, weighted, NHS CHECK cohort, and found no clinically significant differences. This suggests our sample were broadly representative of the NHS workforce in terms of the distress experienced.

Through being independent researchers, we may have facilitated more honest and open communication about services with participants who may have been concerned about confidentiality, than had this research been carried out ‘in-house’ within Trusts. With a mix of clinical and non-clinical interviewers, we were positioned as a mixture of insider and outsider researchers (Dwyer & Buckle, 2009); we attempted to mitigate potential biases in interviewing and data analysis resulting from our respective positions through reflexive discussion throughout the interview and analysis process. We acknowledge that there was a lack of diversity in ethnicity and gender amongst the interviewers (with all being female and White British), and the wider research team (with the majority being White British), which could restrict the breadth of our perception and understanding as well as impact on rapport and data collection in interviews.

### Implications for practice and research

At a socio-political level, and at the risk of stating the obvious, we cannot help but observe that critical workforce shortages that prevent staff from being able to access support continue to contribute to perceptions of an unsupportive workplace. Protected time to access services, breaks and rest periods, while known to be protective of wellbeing are only possible where staff are not covering multiple absences. As many others have pointed out, long-term strategic planning is required to evaluate support services on offer to staff, and to sustain and prioritise the services that are most helpful to staff.

At the organisational level, our findings suggest that staff feel more supported in general, and more able to access specific services, where there is compassionate leadership, supportive line management, and peer support. Line managers in turn require training, resources and flexibility to support their teams, and leaders at all levels should be supported to create virtuous cycles throughout organisations. Frequent reminders and memorable single points of contact for support services are beneficial in raising awareness and simplifying access to support services. Trusts should be aware that some staff feel in-house services are preferrable, but some are wary of stigma around mental health, and so are more comfortable with externally provided services, and so a mix of options is advised.

Our findings indicate that further research in several areas is important. It is important to compare HCW perceptions of workplace support with that of staff who have developed or facilitated these support services to understand whether there is any mismatch between perceived and actual needs for staff. Future research should also expand on findings in this research about how to remove barriers to accessing support, and how the wider socio-political context within which HCWs operate impacts on accessing and using support. Services should be evaluated for effectiveness, accounting for how staff perceive their usefulness and accessibility.

## Conclusion

Our research indicates that enabling positive and caring workplace cultures in healthcare settings, alongside clearly communicated, easily accessible structured support offers, are likely to promote better psychological outcomes for staff and help retain workforce. However, cultural change will be challenging for NHS Trusts without action to address longstanding workforce shortages and system pressures. Such change is necessary to address the underlying causes of psychological distress, as well as reducing the most significant individual barriers to accessing support, i.e. heavy workloads and lack of time. It is not enough to simply provide support services; conditions must be created that enable staff to use them.

## Data Availability

船The data that support the findings of this study are available from the corresponding author (DL), upon reasonable request. The data have not been made publicly available due to the personal and sensitive content of participants experiences.

## Acknowledgements

We wish to acknowledge the National Institute of Health Research (NIHR) Applied Research Collaboration (ARC) National NHS and Social Care Workforce Group, with the following ARCs: East Midlands, East of England, South West Peninsula, South London, West, North West Coast, Yorkshire and Humber, and North East and North Cumbria. They enabled the set-up of the national network of participating hospital sites and aided the research team to recruit effectively during the COVID-19 pandemic. This paper is independent research supported by the National Institute for Health and Care Research ARC North Thames. The views expressed in this publication are those of the author(s) and not necessarily those of the National Institute for Health and Care Research or the Department of Health and Social Care.

The NHS CHECK consortium includes the following site leads: Sean Cross, Amy Dewar, Chris Dickens, Frances Farnworth, Adam Gordon, Charles Goss, Jessica Harvey, Nusrat Husain, Peter Jones, Damien Longson, Richard Morriss, Jesus Perez, Mark Pietroni, Ian Smith, Tayyeb Tahir, Peter Trigwell, Jeremy Turner, Julian Walker, Scott Weich, Ashley Wilkie.

The NHS CHECK consortium includes the following co-investigators and collaborators: Peter Aitken, Anthony David, Sarah Dorrington, Rosie Duncan, Cerisse Gunasinghe, Stephani Hatch, Daniel Leightley, Isabel McMullen, Martin Parsons, Paul Moran, Dominic Murphy, Catherine Polling, Alexandra Pollitt, Danai Serfioti, Chloe Simela, Charlotte Wilson Jones.

